# Effect of Increased Alcohol Consumption During COVID-19 Pandemic on Alcohol-related Liver Disease: A Modelling Study

**DOI:** 10.1101/2021.03.18.21253887

**Authors:** Jovan Julien, Turgay Ayer, Elliot B Tapper, Carolina Barbosa, William Dowd, Jagpreet Chhatwal

## Abstract

**Objectives:** The burden of alcohol-related liver disease (ALD) is surging in the US. There is evidence that alcohol consumption increased during the early periods of the coronavirus disease-2019 (COVID-19) pandemic. We describe the impact of increased alcohol consumption on alcohol-related liver disease.

**Design:** Microsimulation model

**Setting:** Model parameters estimated from publicly available data sources, including national surveys on drug and alcohol use and published studies informing the impact of alcohol consumption on ALD severity.

**Participants:** US residents

**Methods:** We extended a previously validated microsimulation model that estimated the short- and long-term effect of increased drinking during the COVID-19 pandemic in individuals in the US born between 1950-2012. We modelled short- and long-term outcomes of current drinking patterns during COVID-19 (status quo) using survey data of changes in alcohol consumption in a nationally representative sample between February and April 2020. We compared these outcomes with a counter-factual scenario wherein no COVID-19 occurs, and drinking patterns do not change. Reported outcomes are for individuals aged 18-65.

**Main outcome measures:** ALD-related deaths inclusive of HCC mortality, the prevalence and incidence of decompensated cirrhosis and hepatocellular carcinoma, and disability-adjusted life-years (DALYs)

**Results:** Increases in alcohol consumption during 2020 due to the COVID-19 pandemic are estimated to result in to 8,200 [95% UI 7,700 – 8,700] additional ALD-related deaths (1% increase compared with the counter-factual scenario), 17,100 [95% UI 16,100 – 18,200] cases of decompensated cirrhosis (2% increase) and 1,100 [95% UI 1,100 – 1,200] cases of HCC (1% increase) between 2020 and 2040. Between 2020 and 2023, alcohol consumption changes due to COVID-19 will lead to 100 [100-200] additional deaths and 2,200 [2,200-2,300] additional decompensations in patients suffering from alcohol-related liver disease.

**Conclusions:** A short-term increase in alcohol consumption during the COVID-19 pandemic can substantially increase long-term ALD-related morbidity and mortality. Our findings highlight the need for individuals and policymakers to make informed decisions to mitigate the impact of high-risk alcohol drinking in the US.

**Summary Box”:** *What is already known on this topic:*

- The impact of an increase in alcohol consumption during coronavirus disease 2019 (COVID-19) on drinking trajectory changes and alcohol-related liver diseases is not known.
- Studies have reported increases in hospital admissions for alcohol-related liver disease or pancreatitis potentially related to COVID-19, increases in alcohol consumption, and exacerbation of pre-existing liver injury, though limited evidence exists for the long-term effect of increased drinking on alcohol-related liver cirrhosis and liver cancer in the USA.

*Added value of this study:*

- Our study provides new data on liver disease morbidity and mortality associated with increased consumption of alcohol during the COVID-19 pandemic.
- Our study suggests that drinking changes associated with the COVID-19 pandemic it is expected to lead to increases in both mortality and morbidity in the long term. to 8,200 additional ALD-related deaths, 17,100 cases of decompensated cirrhosis, and 1,100 cases of HCC between 2020 and 2040 2

## INTRODUCTION

Since the start of coronavirus disease 2019 (COVID-19) pandemic response in the United States, alcohol consumption has increased considerably -- observed increases in consumption of up to 25% have been recorded in small samples.^1 2^ Weekly sales of alcohol have also been observed to increase by as much as 400% and to be sustained over many weeks.^3 4^ A recent nationally representative survey showed considerable increases in alcohol consumption overall and by populations of interest including females, Black people, and households with children.^1^ There is risk that these short-term increases may manifest as long-term trends in drinking at the societal level. There is an urgent need to understand the impact of these changes and to inform the decisions of individuals, policymakers, and healthcare workers.

The burden of alcohol-related liver disease (ALD) has risen in the past decade, particularly among the young and women. ^5-7^ Short-term increases in alcohol consumption can result in abrupt rises in alcohol use disorder (AUD) and other long-term changes in behavior and care,^8^ which may cause complications such as injury and cirrhosis, leading to long-term increase in ALD and mortality.

As the COVID-19 pandemic continues to evolve, it is important to understand the immediate and long-term effects of increased alcohol consumption on ALD to inform appropriate clinical and policy management. The objective of our study was to quantify projected ALD mortality in the context of increases in alcohol consumption during the COVID-19 pandemic and compare outcomes to a counter-factual scenario without COVID-19 and no changes in drinking patterns.

## METHODS

### Model Overview

We extended a previously developed microsimulation model of fibrosis progression in ALD for US adults^5^ by accounting for current age-cohort and sex-based drinking rates as collected by the National Epidemiologic Survey on Alcohol and Related Conditions (NESARC)^9^, drinking trajectories in the US population^10^, and fibrosis progression rates for drinkers of varying levels that account for current age-cohort and sex-based drinking trajectories in the US population.^11^ At any given time, a patient occupies one of the health states in the model, which is defined by a combination of their liver health stage and drinking level. (Figure 1) Individuals transition from one health state to another based on progression and regression rates that are dependent on drinking level and sex. The model accounts for competing causes of mortality, both related and unrelated to alcohol use (not shown in figure 1 for simplicity). Time advances in one-year increments. The model was developed in R, version 3.6.1 (R Foundation for Statistical Computing) using microsimulation code adapted from DARTH group.^12^

**Figure 1:**
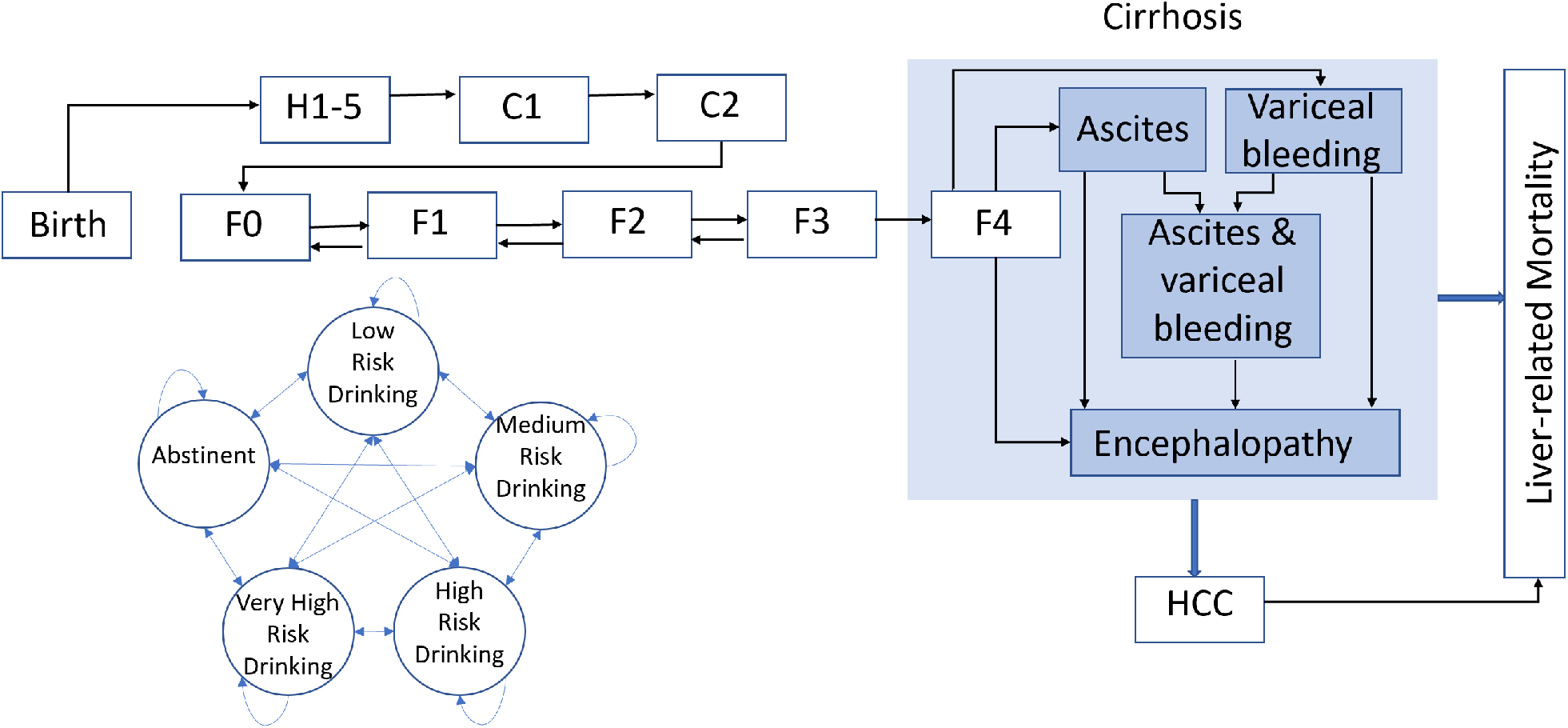
State-transition model of the natural history of alcohol-related liver disease and drinking state. A patient is represented by the combination of one of the health states and one drinking state, shown here as rectangles and circles. Arrows between states represent annual transition probabilities, with the blue shaded states representing decompensated cirrhosis. Competing-cause mortality, the probability of dying from other causes both related and unrelated to alcohol use, exists in every state, but are not shown in our diagram for simplicity. F0, F1, and F2 represent no fibrosis (F0), mild fibrosis (F1) moderate fibrosis (F2). F3 indicates septal fibrosis, F4 compensated fibrosis, and the darker blue stages represent various complications of decompensated fibrosis. The lighter shade of blue encompasses all the cirrhotic disease states. HCC indicates a patient has hepatocellular carcinoma, and mortality from stages F4 on contribute to the reported liver-related mortality.

### Data Sources

Mortality data were collected from the CDC WONDER database for the years of interest.^13^ The queries were placed for ICD-10 codes K70.0 (Alcoholic fatty liver), K70.1 (Alcoholic hepatitis), K70.2 (alcoholic fibrosis and sclerosis of liver), K70.3 (Alcoholic cirrhosis of liver), K70.4 (Alcoholic hepatic failure), K70.9 Alcoholic liver disease following procedures established for National Institute on Alcohol Abuse and Alcoholism surveillance reports on liver cirrhosis mortality in the United States.^14^ Other natural history transition probabilities are sourced from existing research as outlined above. (Supplement tables 1 and 2)

We estimated individual drinking levels using the NESARC studies,^9^ limited access datasets obtained from the National Institute on Alcohol Abuse and Alcoholism, which are nationally representative surveys of at least 36,309 US adults collected between 2001-2002 (NESARC-I Wave 1) and April 2012 and June 2013 (NESARC-III). The survey data allows for estimates of the 2001-2002 and 2012-2013 distribution of drinkers by age and gender. Compared against data collected 10 years prior during NESARC-I, the NESARC-III data also allowed for the consideration of changing rates of high-risk drinking amongst age groups.^9^

In May 2020, members of our research team conducted a survey of 993 individuals using a probability-based panel designed to be representative of the US population aged 21 and older. We found that average drinks per day and excessive drinking (drinking in excess of recommended drinking limits and binge drinking) increased significantly by 29% and 21% between February (before stay-at-home orders were issued) and April (when stay-at-home orders were in place in many states).^1^

### Population

We model birth cohorts of US adults from 1950-2012. To capture an approximation of drinking before 2012, individuals are assigned a drinking level from the 2012 distribution collected in NESARC-III for their age group and allowed to progress until 2012. In 2012, drinking rates returned to the initial level and progress again according to the drinking transition matrix.

### Drinking levels

Drinking levels consist of never yet consumed alcohol (denoted H1 – H5 in the model), abstinence, low risk (< 3 drinks per day), medium risk (3-4) drinks per day), high risk (4-7 drinks per day), and very high risk states (> 7 drinks per day). Transitions are possible between every drinking level (abstinence, low risk, medium risk, high risk, and very high risk drinking) and transition rates are consistent with rates found in Barbosa et al., 2019.^10^

We calibrate transition probabilities in the tunnel states H1-H5 such that the distribution of simulated individuals’ initiation of drinking, the age at which they transition out of H5, matched initiation of drinking data in NESARC-III data.

### Natural history of ALD

The following define the liver-related health states: no fibrosis (F0), mild fibrosis (F1), moderate fibrosis (F2), and septal fibrosis (F3). Transition probabilities between states F1-F3 replicate fibrosis progression and regression in Lieber et al. 2003 and are responsive to changes in drinking rate.^11^ Liver-related health is defined by the following stages: no fibrosis (F0), mild fibrosis (F1), moderate fibrosis (F2), and septal fibrosis (F3). Transition probabilities between states F1-F3 replicate fibrosis progression and regression in Lieber et al. 2003 and are responsive to changes in drinking rate.^11^

Patients can further progress to compensated cirrhosis (F4), decompensated cirrhosis (ascites, variceal bleeding, ascites with variceal bleeding, and encephalopathy), as well as hepatocellular carcinoma (HCC). From these advanced disease stages, patients can die from liver-related mortality. Real-world transition probabilities are used to estimate transition probabilities between compensated cirrhosis (F4), decompensated cirrhosis (ascites, variceal bleeding, ascites with variceal bleeding, and encephalopathy), and death consistent with the findings of Jepsen et al.^15^ Mortality rates from advanced stages of liver disease are estimated from population studies that allowed for transplantation; while a transplantation state is not explicitly delineated in our model, we account for the competing event of transplantation before death.^15^ Health state transitions were dependent on progression and regression rates associated with different levels of drinking.^11^ All transition probabilities are summarized in Supplement tables 1, 2, and 3.

States C1 and C2 are calibration states used to capture unidentified interactions, including genetic, age-cohort, gender, diet, and other factors known to influence the progression of liver disease but not explicitly captured in the model. We calibrate the model overall by adjusting the C1 and C2 transition probabilities to replicate observed mortality data from the CDC WONDER database for 2012-2014.^13^ Individuals born after 1980 were calibrated as a group and for a longer observation period (2010 – 2014) due to lack of mortality data.

### Simulated scenarios

To project model outcomes up to 2040, we consider two scenarios for drinking in the US population. (1) ‘COVID-19 consumption scenario’ describes a measured increase in alcohol consumption based on recorded increases in the Barbosa et al. study of individuals (n = 993) in the US population described earlier in the manuscript.^16^ (2) The ‘counter-factual scenario’ describes the absence of COVID-19 related effects. This scenario assumed that age-specific rates of alcohol consumption continue to evolve in the absence of COVID-19.

### Model outcomes

We project ALD-related deaths inclusive of HCC mortality, the prevalence and incidence of decompensated cirrhosis and HCC, and disability-adjusted life-years (DALYs) lost to ALD from 2020 to 2040. DALYs — a combined measure of disability, prevalence of disease (in our case decompensated cirrhosis and HCC), and quantity of life-years lost due to disease and mortality^34-36^— were calculated using disability weights from 2016 Global Burden of Disease study values for decompensated cirrhosis and HCC.^37^ Life-years lost due to disease and mortality (YLL) were estimated by multiplying each death by the remaining life expectancy at the age of death and summing across the years. To calculate years lived with disability (YLD), we multiply the number of individuals in a health state by the disability weights defined by the Global Burden of Disease study (0 for scores of F0–F4, 0.178 for decompensated cirrhosis, and 0.540 for HCC).^38,39^ DALYs are then the sum of YLLs and YLDs.

### Sensitivity analysis

We conducted a probabilistic sensitivity analysis to determine the confidence in our model projections in each scenario given the joint uncertainty of model parameters, in particular natural history of disease, transition rates, and drinking distribution. For this purpose, we first defined parameter uncertainty using recommended distributions for different types of parameters (see **Supplement Tables 2**).^17^ We then sampled parameter values from these distributions 1000 times and ran the model to determine model results. The populations used were 1/10^th^ the size of the US birth cohort. We generated the 95% uncertainty intervals of model outcomes.

## RESULTS

We first validate our model-predicted annual mortality due to alcohol-related liver disease from 2014 to 2018 for men and women with those reported by the US national death registry data from the CDC WONDER database **(Figure 2)**. We also compare alcohol-related cirrhosis prevalence in the US population in our model against reported data in 2009 and 2015 in the peer reviewed literature (**Supplement Table 5)**.^18^

**Figure 2:**
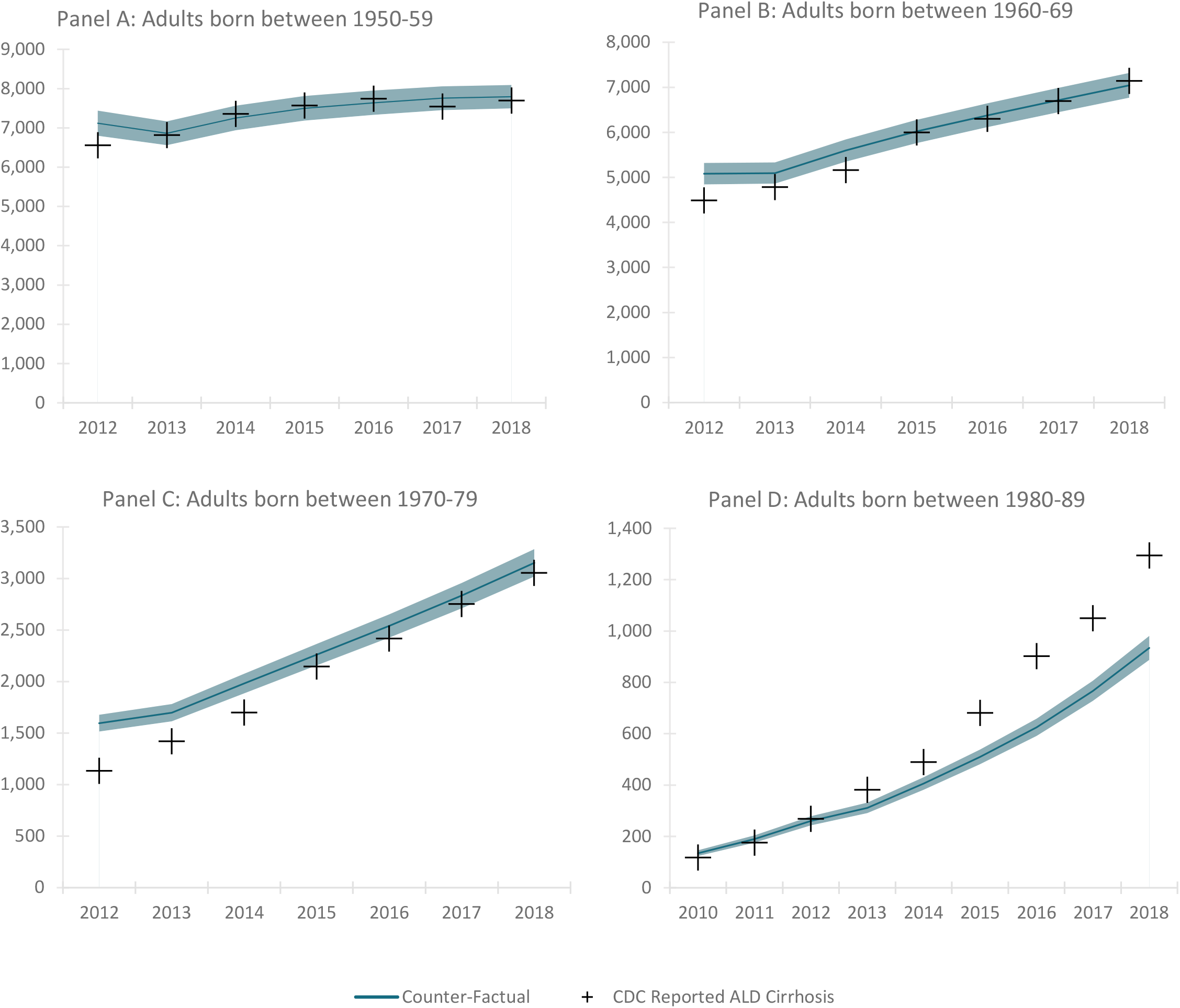
Birth cohort ALD disease mortality 2012-2018 with CDC mortality validation. Mortality as reported in the US National Death Registry and predicted/projected mortality in our model from the start of the calibration period through the end of the validation period. Shaded regions represent the confidence intervals generated by probabilistic sensitivity analysis.

Under the counter-factual scenario, which assumed no COVID-19 associated drinking changes, 655,300 [95% uncertainty interval (UI) 628,500-682,200] US adults, aged 18-65, are projected to die from ALD-related cirrhosis between 2020 and 2040, 830,500 [95% UI 793,200 – 867,800] are projected to develop decompensated cirrhosis, and 104,900 [95% UI 100,000 - 109,800] are projected to develop HCC during the same period **(Table 1)**.

**Table 1:** Cumulative projections for ALD disease morbidity and mortality for 3- and 20-year horizons under COVID-19 consumption scenario and counter-factual scenario. The table presents cumulative alcohol-related liver disease (ALD) morbidity and mortality for two time periods, 2020-2023 and 2020-2040. Data is presented as a the mean of model results and the 95% confidence interval generated by the probabilistic sensitivity analysis. Morbidity results include the incidence of decompensated cirrhosis and hepatocellular carcinoma, two diseases with significant health implications. In the COVID-19 consumption scenario some drinkers increase consumption during 2020 while in the counter-factual drinking progresses without any increases associated with COVID-19.

| Scenario | Results | Short term horizon<br>(2020 – 2023) | Long term horizon<br>(2020 – 2040) |
| --- | --- | --- | --- |
| COVID-19<br>Consumption | *ALD Cirrhosis Mortality* | 83,100<br>[79,700-86,500] | 663,500<br>[636,400-690,700] |
|  | *Decompensated Cirrhosis Incidence* | 66,500<br>[63,700-69,300] | 847,700<br>[809,300-886,100] |
|  | *HCC Incidence* | 13,300<br>[12,600-13,900] | 106,000<br>[101,100-111,000] |
| Counter-factual | *ALD Cirrhosis Mortality* | 83,000<br>[79,600-86,300] | 655,300<br>[628,700-682,000] |
|  | *Decompensated Cirrhosis Incidence* | 64,300<br>[61,500-67,000] | 830,500<br>[793,200-867,800] |
|  | *Hepatocellular Carcinoma Incidence* | 13,300<br>[12,700-13,900] | 104,900<br>[100,000-109,800] |
| Increase in outcomes<br>because of COVID-19<br>drinking | *ALD Cirrhosis Mortality* | 100<br>[100-200] | 8,200<br>[7,700-8,700] |
|  | *Decompensated Cirrhosis Incidence* | 2,200<br>[2,200-2,300] | 17,100<br>[16,100-18,200] |
|  | *Hepatocellular Carcinoma Incidence* | 00<br>[00-00] | 1,100<br>[1,100-1,200] |
|  | *DALYs* | 198,400<br>[194,500 - 202,300] | 1,675,600<br>[1,668,900-1,682,400] |

Under the ‘COVID-19 consumption scenario’, 663,500 [95% UI 636,400-690,700] US adults, aged 18-65, are projected to die from ALD cirrhosis between 2020 and 2040, 847,700 [95% UI 809,300 886,100] are projected to develop decompensated cirrhosis, and 106,000 [95% UI 101,100 - 111,000] are projected to develop HCC **(Table 1)**. Increase in alcohol consumption due to COVID-19 in 2020 is projected to lead to 8,200 [95% UI 7,700 – 8,700] additional deaths, 17,100 [95% UI 16,100 – 18,200] additional decompensation cases and 1,100 [95% UI 1,100 – 1,200] additional cases of HCC over the next 2 decades **(Figure 3)**. We also estimated a pronounced spike in decompensation cases and mortality in the short term, during 2020-2021.

**Figure 3.**
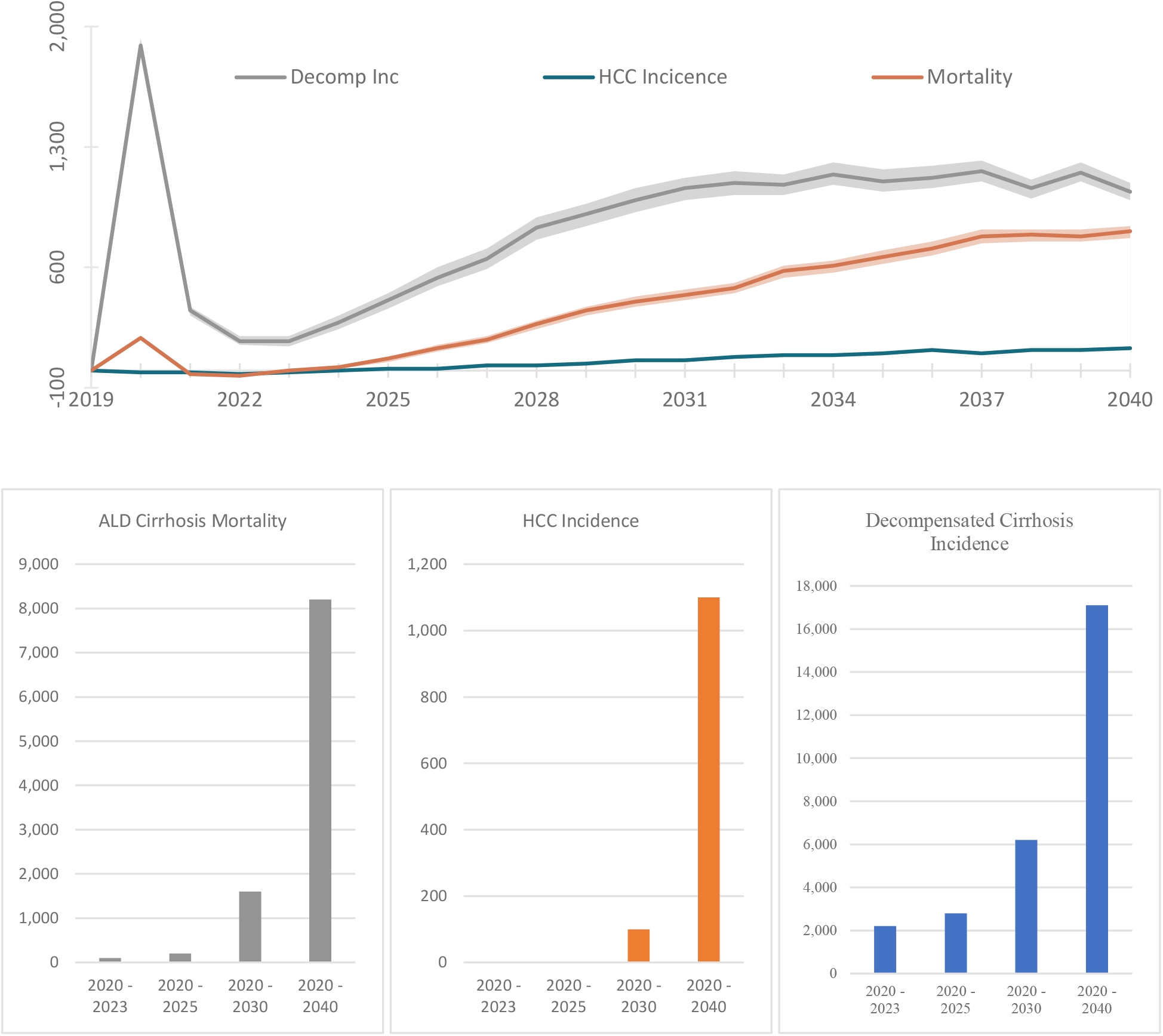
Per-annum and cumulative difference in alcohol-related liver mortality and morbidity between COVID-19 consumption scenario and counter-factual for USA adults aged 18-65, 2019-2040. Annual and cumulative rates for morbidity and mortality differences between the COVID-19 consumption scenario and counter-factual scenario associated with alcohol-related liver disease. In the COVID-19 consumption scenario some drinkers increase consumption during 2020 during the pandemic while in the counter-factual drinking progresses without any increases associated with COVID-19. Shaded regions represent the 95% confidence interval associated with the probabilistic sensitivity analysis.

### Age Cohort Related Mortality

Alongside understanding increases in morbidity and mortality generally, we sought to explore the impact on specific age groups. For those born in the 1950s, the model projects approximately 142,810 deaths from 2020-2040 in the ‘COVID-19 consumption scenario’ vs 142,210 deaths in the ‘counter-factual scenario’, our projections indicate that for the birth cohort born in the 1970s we expect 235,040 and 232,730 deaths per year respectively. While the initial mortality increases are concentrated in the 1950s birth cohort, over the next 20 years the additional ALD mortality burden shifts increasingly to the younger birth cohorts **(Table 2)**.

**Table 2:**
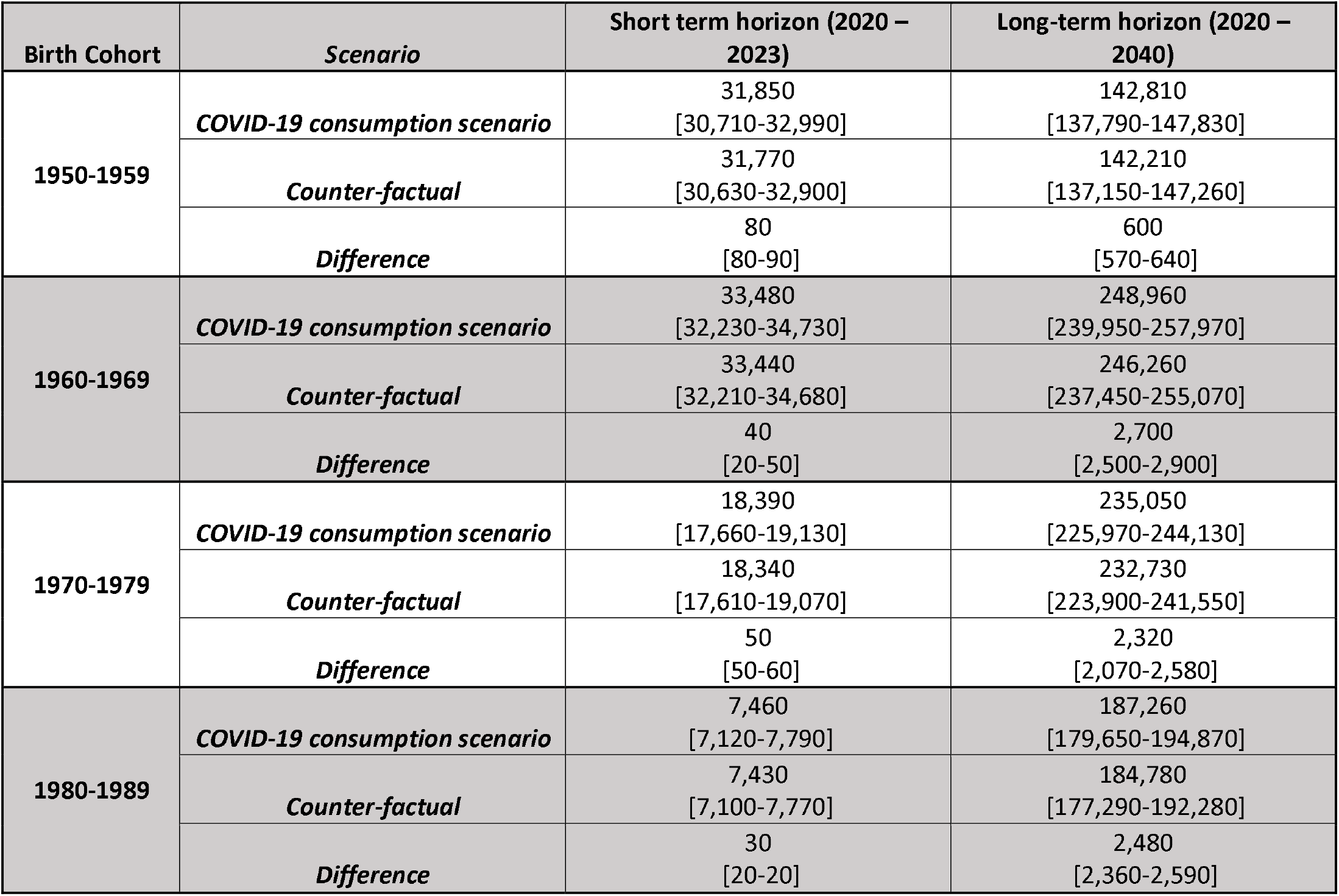
Cumulative projections for ALD disease mortality by birth cohort for 3and 20-year horizons under COVID-19 consumption scenario and counter-factual scenario.

### Disability Adjusted Life-Years

COVID-19 related drinking increases are also associated with substantial DALYs, a measure of both life years lost due to premature mortality and years of productive life lost due to disability. From 2020-2023, drinking increases during COVID-19 are projected to lead to 198,400 DALYs relative to the status-quo, a number which is projected to increase to 1,675,600 cumulative DALYs from 2020 through 2040 (**Table 1**).

## DISCUSSION

Alcohol-related liver disease accounts for a significant health burden in the United States.^5^ Alcohol consumption has increased substantially during the COVID pandemic.^1 2^ Our results show that there may be a measurable effect on ALD-related decompensation and mortality in the short-term, and, without intervention, it can result in a long-term disease burden of ALD-related HCC and mortality. The observed increase in alcohol consumption has the potential to exasperate the condition of those already living with impaired liver function and to lead to long-term damage in those who increase consumption at an early age.

### ALD is a major threat to public heath

ALD already poses a significant disease burden in the United States. ALD now accounts for more than $5 billion in direct healthcare costs and mortality due to ALD has increased substantially, by 200% in people aged 25-34 over the last decade. ^18 19^ Even without COVID related increases in consumption, ALD will claim the lives of nearly 260,000 Americans over the next decade. Our models show that if the higher rates of risky alcohol consumption hold for one year, this burden will further rise. A key insight from these data, however, is that in the short-term there will be a disproportionate rise in the excess morbidity and mortality attributed to ALD. Many with ALD, even those with compensated cirrhosis, are clinically stable but marginal increases in consumption for these vulnerable patients are uniquely toxic.^20^ The result, we find, is a spike in the short-term incidence of decompensated cirrhosis that will stress a healthcare system that is still overwhelmed by the impact of the pandemic.^8^

### The burden of ALD requires a public health approach

Our data shows that COVID-associated increases in alcohol consumption increase short- and long-term morbidity and mortality due to ALD. However, even in the counter factual estimates, we expect a steady expansion of the public health impact of ALD. Our data shows also that young persons will be disproportionately impacted. There is, accordingly, substantial value in interventions to reduce the burden of ALD. All ALD-focused interventions aim to reducing alcohol consumption. At the individual level, one can use behavioral and pharmacological interventions, each proven to reduce the harms of alcohol use disorder.^21^ However, the impact of these interventions remains limited owing to modest efficacy and inadequate linkage to care.^22 23^ Our data underscores a population-level problem for which a population-level intervention may be indicated. Policies that reduce access to alcohol especially during the COVID-19 pandemic or influence demand through minimum unit pricing may be the most effective tools.^24-27^ In Scotland, a minimal unit price of 50 pence has resulted in a substantial decrease in alcohol consumption, primarily by the highest consumers.^28^

### Contextual factors

Our results must be interpreted in the context of the study design. First, there is a lack of comprehensive information on the increase, and distribution of increase, of drinking in the US population. Accordingly, we utilize data from a representative sample of the US population taken in 2020 as a conservative representation of the impact of drinking increases. To mitigate this limitation, we model the uncertainty in our estimates with extensive and conservative probabilistic sensitivity analysis. Second, there are other (unaccounted) factors that could impact the risk of ALD-related decompensation. The significant turmoil of this moment in the United States caused by COVID-19, and the associated economic and political stressors, may have had disproportionate impact on individuals at increased risk of adverse health outcomes.^29^ Individuals often increase alcohol intake to cope with emotional stress and chronic uncertainty; these coping mechanism have unclear long-term consistency^30^. To address this concern, our increased consumption scenario only increases drinking for one year whereupon drinking transitions return to the pre-2020. Third, this modelling study does not account for the impact of COVID-19 infection on those with pre-existing cirrhosis or those at-risk for future decompensation. Fourth, we do not model the impact of adverse trends on alcohol consumption on specific population subgroups. Given recent complementary results by Barbosa et al.,^16^ a particular focus on at risk groups including females, Black people, and households with children, should be taken as increased morbidity and mortality beyond the status quo should be expected to be concentrated in populations with the greatest increase in consumption and fibrosis susceptibility. While the study does not specifically address the impacts in at risk communities, the general dynamics of consumption, liver damage, and morbidity and mortality described here should remain similar.

### Conclusion

Our study raises the important specter of the near- and long-term impact of alcohol-related liver disease. Even a short-term increase in alcohol consumption during the pandemic can substantially increase morbidity and mortality associated with ALD in the long term due to behavioral changes. Our findings highlight the need for individuals and policy makers to make informed decisions to mitigate the impact of high-risk alcohol drinking while staying at home during the COVID-19 pandemic in the US. Short-term increases in consumption are associated with both an immediate bump in mortality and morbidity, as well as long term increases many years later.

## Data Availability

Data are available.

## ACKNOWLEDGMENTS

1. Jagpreet Chhatwal is the guarantor of this article
2. Roles
  a. Concept: Jovan Julien and Jagpreet Chhatwal
  b. Analysis: Jovan Julien
  c. Data acquisition: Jovan Julien, Carolina Barbosa, William Dowd
  d. Analysis and interpretation of data: Jovan Julien, Turgay Ayer, Elliot Tapper, Jagpreet Chhatwal
  e. Writing: Jovan Julien, Jagpreet Chhatwal
  f. Revision: Turgay Ayer, Elliot Tapper, Carolina Barbosa, William Dowd, Jagpreet Chhatwal

## 5. Ethics approval

The study (Protocol H18459) obtained ethics approval through the institutional review board of Georgia Institute of Technology.

## 6. Transparency Statement

Jagpreet Chhatwal affirms that this manuscript is an honest, accurate, and transparent account of the study being reported; that no important aspects of the study have been omitted; and that any discrepancies from the study as planned (and, if relevant, registered) have been explained.

## References

1. Barbosa C, Cowell AJ, Dowd WN. Alcohol Consumption in Response to the COVID-19 Pandemic in the United States. Journal of Addiction Medicine 2020

2. Pollard MS, Tucker JS, Green HD. Changes in adult alcohol use and consequences during the COVID-19 pandemic in the US. JAMA network open 2020;3(9):e2022942–e42.

3. IZEA. Coronavirus Impacts on Alcohol & Social Media Consumption. IZEA. IZEA Insights, 2020.

4. Drizly.com. How alcohol ecommerce sales are being impacted across North America, 2020.

5. Julien J, Ayer T, Bethea ED, et al. Projected prevalence and mortality associated with alcohol-related liver disease in the USA, 2019–40: a modelling study. The Lancet Public Health 2020;5(6):e316–e23.

6. Tapper EB, Parikh ND. Mortality due to cirrhosis and liver cancer in the United States, 1999-2016: observational study. BMJ 2018;362 doi: 10.1136/bmj.k2817

7. White AM, Castle IJP, Hingson RW, et al. Using death certificates to explore changes in alcohol-related mortality in the United States, 1999 to 2017. Alcoholism: Clinical and Experimental Research 2020;44(1):178–87.

8. Tapper EB, Asrani SK. COVID-19 pandemic will have a long-lasting impact on the quality of cirrhosis care. Journal of hepatology 2020

9. Grant B, Chu A, Sigman R, et al. Source and accuracy statement: national epidemiologic survey on alcohol and related conditions-III (NESARC-III). Rockville, MD: National Institute on Alcohol Abuse and Alcoholism 2014:1–125.

10. Barbosa C, Dowd WN, Aldridge AP, et al. Estimating long-term drinking patterns for people with lifetime alcohol use disorder. Medical decision making 2019;39(7):765–80.

11. Lieber CS, Weiss DG, Groszmann R, et al. I. Veterans Affairs Cooperative Study of polyenylphosphatidylcholine in alcoholic liver disease: effects on drinking behavior by nurse/physician teams. Alcoholism: Clinical and Experimental Research 2003;27(11):1757–64.

12. Krijkamp EM, Alarid-Escudero F, Enns EA, et al. Microsimulation Modeling for Health Decision Sciences Using R: A Tutorial. Med Decis Making 2018;38(3):400–22. doi: 10.1177/0272989X18754513

13. Centers for Disease Control and Prevention NCfHS. Underlying Cause of Death 1999-2019 on CDC WONDER Online Database, released in 2020. Data are from the Multiple Cause of Death Files, 1999-2019, as compiled from data provided by the 57 vital statistics jurisdictions through the Vital Statistics Cooperative Program. 2020 [Available from: http://wonder.cdc.gov/ucd-icd10.html accessed April 8 2020.

14. Yoon Y-H, Chen CM. Surveillance Report# 105: Liver cirrhosis mortality in the United States: National, state, and regional trends, 2000–2013. National Institute on Alcohol Abuse and Alcoholism (NIAAA), Bethesda, MD 2016

15. Jepsen P, Ott P, Andersen PK, et al. Clinical course of alcoholic liver cirrhosis: A Danish population-based cohort study. Hepatology 2010;51(5):1675–82.

16. Barbosa C, Cowell, A, Dowd, W.,. How Has Drinking Behavior Changed During the COVID-19 Pandemic? Results from a Nationally Representative survey. : RTI International.

17. Briggs AH, Weinstein MC, Fenwick EAL, et al. Model Parameter Estimation and Uncertainty Analysis: A Report of the ISPOR-SMDM Modeling Good Research Practices Task Force Working Group–6. Medical Decision Making 2012;32(5):722–32. doi: 10.1177/0272989X12458348

18. Mellinger JL, Shedden K, Winder GS, et al. The high burden of alcoholic cirrhosis in privately insured persons in the United States. Hepatology 2018;68(3):872–82. doi: 10.1002/hep.29887

19. Tapper EB, Parikh ND. Mortality due to cirrhosis and liver cancer in the United States, 1999-2016: observational study. BMJ 2018;362:k2817.

20. Lucey MR, Connor JT, Boyer TD, et al. Alcohol consumption by cirrhotic subjects: patterns of use and effects on liver function. American Journal of Gastroenterology 2008;103(7):1698–706.

21. Peng JL, Patel MP, McGee B, et al. Management of alcohol misuse in patients with liver diseases. Journal of Investigative Medicine 2017;65(3):673–80.

22. Addolorato G, Mirijello A, Barrio P, et al. Treatment of alcohol use disorders in patients with alcoholic liver disease. J Hepatol 2016;65(3):618–30. doi: 10.1016/j.jhep.2016.04.029

23. Rogal S, Youk A, Zhang H, et al. Impact of alcohol use disorder treatment on clinical outcomes among patients with cirrhosis. Hepatology 2020;71(6):2080–92.

24. Xuan Z, Blanchette JG, Nelson TF, et al. Youth drinking in the United States: Relationships with alcohol policies and adult drinking. Pediatrics 2015;136(1):18–27.

25. Naimi TS, Blanchette J, Nelson TF, et al. A new scale of the U.S. alcohol policy environment and its relationship to binge drinking. Am J Prev Med 2014;46(1):10–6. doi: 10.1016/j.amepre.2013.07.015

26. Black H, Gill J, Chick J. The price of a drink: levels of consumption and price paid per unit of alcohol by Edinburgh’s ill drinkers with a comparison to wider alcohol sales in Scotland. Addiction 2011;106(4):729–36.

27. Holmes J, Meng Y, Meier PS, et al. Effects of minimum unit pricing for alcohol on different income and socioeconomic groups: a modelling study. The Lancet 2014;383(9929):1655–64.

28. O’Donnell A, Anderson P, Jané-Llopis E, et al. Immediate impact of minimum unit pricing on alcohol purchases in Scotland: controlled interrupted time series analysis for 2015-18. bmj 2019;366:5274.

29. Pfefferbaum B, North CS. Mental health and the Covid-19 pandemic. New England Journal of Medicine 2020;383(6):510–12.

30. Esterwood E, Saeed SA. Past epidemics, natural disasters, COVID19, and mental health: learning from history as we Deal with the present and prepare for the future. Psychiatric quarterly 2020:1–13.

