## Appendix for "Effect of Increased Alcohol Consumption During COVID-19 Pandemic on Alcohol-related Liver Disease: A Modelling Study"

### **Supplement. Data and Supporting Results**

This online Appendix accompanies the manuscript entitled, “Effect of Increased Alcohol Consumption During COVID-19 Pandemic on Alcohol-related Liver Disease: A Modelling Study.” The Appendix provides data and supporting results, including additional model information.

**Drinking transitions and initialization of model-**

We model birth cohorts of US adults from 1950-2012. To capture an approximation of drinking before 2012, individuals are assigned a drinking rate from the 2012 distribution for their age and gender group at inception of drinking, leaving H5, and allowed to progress until 2012. In 2012, drinking rates return to the initial rate and progress again according to the drinking transition matrix.

***Relevant Data***

#### **Table 1. 2-year Disease Progression Characteristics of Drinking Population in United States (Perivenular Fibrosis (F1), Septal Fibrosis (F2), Incomplete Cirrhosis (F3), Complete/Compensated Cirrhosis (F4)^20^**

|  | Abstinent | Low Risk  3 drinks / day or less  1-40 g/day^20^ | Medium Risk  3-4 drinks/ day  41-60 g/day^20^ | High Risk  4-7 drinks / day  61-105 g/day | Very High Risk  > 7 drinks / day^20^  >106 g/day^20^ |
| --- | --- | --- | --- | --- | --- |
| Regression |  | 28.9% | 24.2% |  | 18.5% |
| Remain Same |  | 59% | 54.8% |  | 63% |
| Progression |  | 12.1% | 21% |  | 18.5% |
| Interpreted for Use in Model |  |  |  |  |  |
| Regression | 28.9% | 28.9% | 24.2% | **21%** | 18.5% |
| Remain Same | 71.1% | 59% | 54.8% | **58%** | 63% |
| Progression | 0% | 12.1% | 21% | **21%** | **21%** |

#### **Table 2. Input parameters used in the state-transition model.**

| ALD Natural History | Probability | Range | Sample  Distribution for Probabilistic Sensitivity Analysis | Ref |
| --- | --- | --- | --- | --- |
| H_1_ 🡪 H_2_  H_2_ 🡪 H_3_  H_3_ 🡪 H_4_  H_4_ 🡪 H_5_  H_5_ 🡪C_0_ | 4·87% | 3·92% - 4·88% | Uniform | ^71^ |
| F4 🡪 Ascites | 8·37% | 4·57% - 18·25% | Beta | ^36^ |
| F4 🡪 Variceal Bleeding | 4·65% | 2·54% - 10·14% | Beta | ^36^ |
| F4 🡪 Encephalopathy | 2·17% | 1·19% - 4·73% | Beta | ^36^ |
| F4 🡪 Cirrhosis Death | 6·72% | 3·72% - 14·87% | Beta | ^36^ |
| Ascites 🡪 Ascites + Variceal Bleeding | 6·57% | 4·60% - 9·85% | Beta | ^36^ |
| Ascites 🡪 Encephalopathy | 5·97% | 4·18% - 8·96% | Beta | ^36^ |
| Ascites 🡪 Cirrhosis Death | 7·46% | 5·22% - 11·19% | Beta | ^36^ |
| Variceal Bleeding 🡪 Ascites + Variceal Bleeding | 9·90% | 3·44% - 32·29% | Beta | ^36^ |
| Variceal Bleeding 🡪 Encephalopathy | 7·35% | 2·55% - 23·96% | Beta | ^36^ |
| Variceal Bleeding 🡪 Cirrhosis Death | 5·75% | 2·00% - 18·75% | Beta | ^36^ |
| Ascites + Variceal Bleeding 🡪 Encephalopathy | 17·5% | 9·5% - 37·5% | Beta | ^36^ |
| Ascites + Variceal Bleeding 🡪 Cirrhosis Death | 17·5% | 9·5% - 37·5% | Beta | ^36^ |
| Encephalopathy 🡪 Cirrhosis Death | 32% | 27% - 37% | Beta | ^36^ |
| F4, Ascites, Variceal Bleeding, Ascites + Variceal Bleeding, or Encephalophathy 🡪 Hepatocellular Carcinoma | 1·7% | 1·2% - 2·2% | Beta | ^37^ |
| HCC -> Cirrhosis Death | 39% |  | Beta | ^38^ |

#### **Table 3. One Year Transition Probabilities Matrix Derived from Barbosa et al. 2019.**

|  | Abstinence | Low-Risk Drinking | Medium-Risk Drinking | High-Risk Drinking | Very High-Risk Drinking | SUM |
| --- | --- | --- | --- | --- | --- | --- |
| Abstinence | 98.4% | 1.1% | 0.2% | 0.2% | 0.1% | 100.0% |
| Low-Risk Drinking | 1.3% | 95.4% | 0.4% | 1.7% | 1.2% | 100.0% |
| Medium-Risk Drinking | 0.4% | 7.4% | 84.3% | 7.6% | 0.3% | 100.0% |
| High-Risk Drinking | 1.2% | 1.4% | 4.8% | 83.5% | 9.2% | 100.0% |
| Very High-Risk Drinking | 3.0% | 0.4% | 3.9% | 2.1% | 90.6% | 100.0% |
| SUM | 104.4% | 105.6% | 94.5% | 95.0% | 101.4% |  |

#### **Table 4. Population Characteristics of Drinking Population in United States from NESARC**

|  | 18-30 | 31-40 | 41-50 | 51-60 | 61-70 | 71-80 | 80+ | Total |
| --- | --- | --- | --- | --- | --- | --- | --- | --- |
| Abstinent | 1,895 | 1,521 | 1,742 | 1,973 | 1,673 | 1,185 | 661 | 10,650 |
| Low-Risk | 6,217 | 4,767 | 4,294 | 3,764 | 2,415 | 1,084 | 527 | 23,068 |
| Medium-Risk | 287 | 199 | 181 | 174 | 89 | 29 | 12 | 971 |
| High-Risk | 260 | 163 | 166 | 149 | 72 | 20 | 6 | 836 |
| Very High-Risk | 229 | 165 | 192 | 137 | 53 | 5 | 3 | 784 |
| Total | 8,888 | 6,815 | 6,575 | 6,197 | 4,302 | 2,323 | 1,209 | 36,309 |

#### **Table 5. Model validation prevalence rates of alcohol-related cirrhosis: Real World**^[1]^ **vs Model**

| Cohort | Real World  2009 | Real World  2015 | Model  2009 | Model  2015 |
| --- | --- | --- | --- | --- |
| 18-65 w/o HCV | 0.05% | 0.072% | 0.057% | 0.076% |
| Younger than 45 | 0.01% | 0.03% | 0.025% | 0.027% |
| Older than 45 | 0.13% | 0.19% | 0.12% | 0.15% |
| Male | 0.10% | 0.13% | 0.074% | 0.98% |
| Female | 0.04% | 0.06% | 0.039% | 0.054% |

#### **Figure 1. Expanded model with tunnel states described**

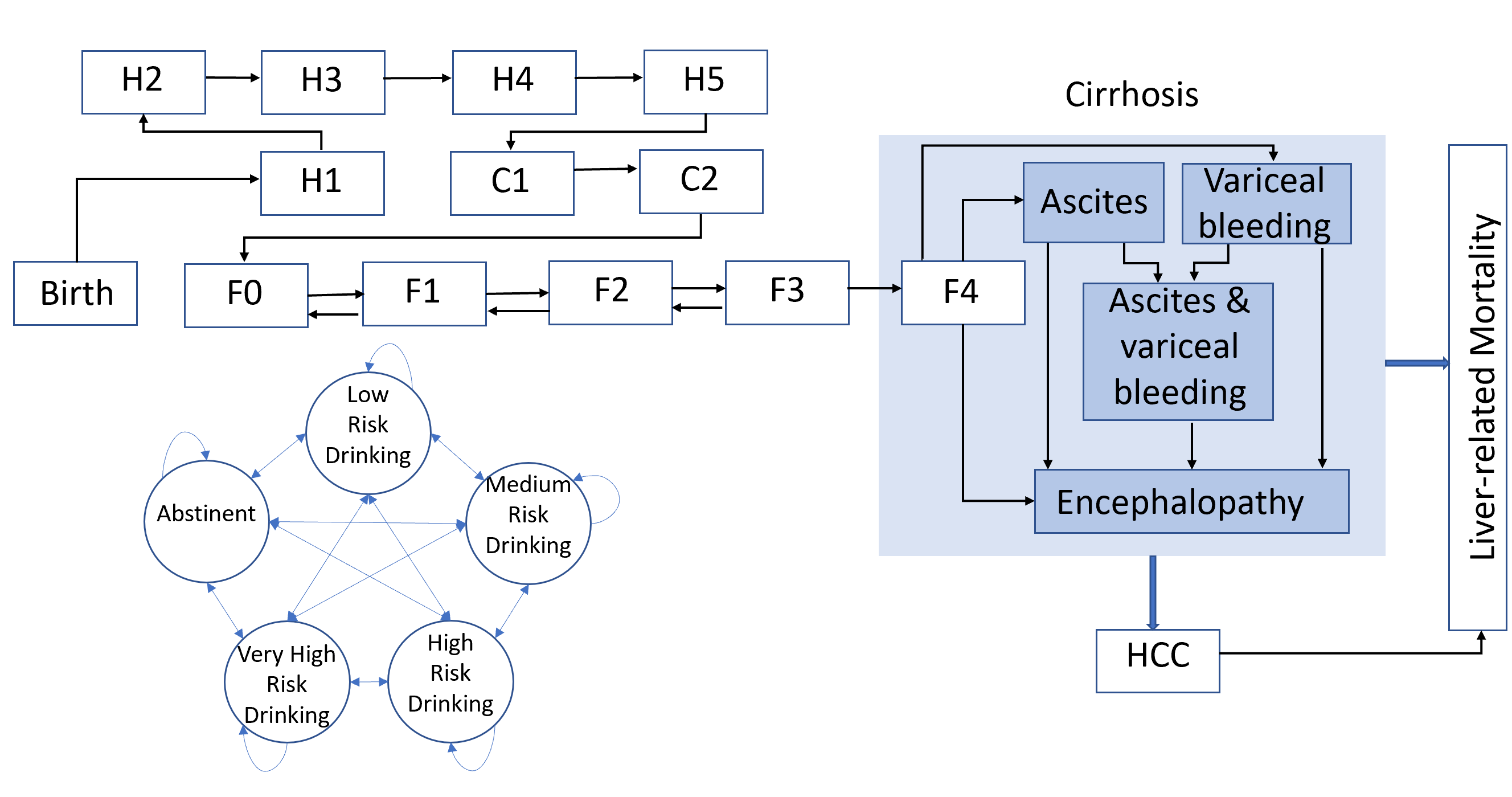
The expanded model shows the details of the tunnel states H1-H5. These states are calibrated to reproduce the age at first drink distributions for the US population as evidenced in the National Epidemiologic Survey on Alcohol and Related Conditions-III.

1. Mellinger JL, Shedden K, Winder GS, et al. The high burden of alcoholic cirrhosis in privately insured persons in the United States. *Hepatology.* 2018;68(3):872-882.
